# Prevalence of Hepatitis B viral infection and associated factors among adults in Moyo district, North-Western Uganda

**DOI:** 10.1101/2022.06.23.22276817

**Authors:** Patrick Madrama Lulu, John Bosco Alege

## Abstract

**Background:** Hepatitis B viral infection is a public health problem and estimates show that about 30% of the world’s population is infected with the virus, with about 350 to 400 million of them remaining chronically infected. Northern Uganda has the highest prevalence of HBV in Uganda. In this study, we sought to establish factors influencing the prevalence of Hepatitis B viral infection among adults in Moyo district, North-Western Uganda.

**Methods:** We used a descriptive cross-sectional study where quantitative data collection methods and analysis was employed. Self-reported HBV infection by respondents who had test result forms diagnosed with HBV within the last one year was reviewed by the researchers to confirm whether the respondent was negative or positive with Hepatitis B. 384 samples were determined using Cochran (1963:75) and a multi-stage sampling technique was used. Bivariate and multivariate analyses were done using SPSS (20.0).

**Results:** Out of the 384 respondents interviewed, 29 (7.6%, 95%CI: 5.1-10.7) had HBV. Factors influencing the prevalence of HBV were; level of education (p=0.047*), ever having had STIs (aOR=18.090, 95%CI=5.699-57.426, p=0.000*), Health facilities have equipment for screening HEP B viral infection ANC (aOR=10.762, 95%CI=1.316-88.027, p=0.027*) were statically significant in influencing the prevalence of HBV, while the number of sexual partners ever had (p=0.984) was not statistically significant in influencing the prevalence of Hepatitis B viral infection.

**Conclusion:** We found a high prevalence of HBV among adults in Moyo district in North-Western Uganda compared to the national prevalence. Key influences of HBV prevalence included education level, ever being infected with any other STIs (HIV, Syphilis, gonorrhoea), multiple sexual partners and presence of HBV screening equipment at the health facilities. Need for more emphasis on HBV childhood immunization, screening, vaccination of adults, other preventive measures and treatment of those already infected.

## Background

Hepatitis B viral (HBV) infection is a major public health problem worldwide and estimates show that about 30% of the world’s population that is close to 2 billion people are infected with the virus, and about 350 to 400 million of them remaining chronically infected and therefore carriers of the virus causing global burden known to be a cause of more than one million deaths [1, 2]. It is the world’s most contagious liver infection more infectious than HIV and the most common ways of transmission are through unprotected sex, unsafe blood transfusion, unsafe use of needles, from mother to child at birth, close household contacts and between children early in childhood [1].

Vaccines for HBV were introduced in 1982 and was integrated into the EPI, it was expected to give 90-100% protection against the infection, but there is still a global challenge where more than 350 million are living with chronic Hepatitis B and approximately 600,000 HBV related deaths per year globally [1], with the most unrelenting health problem found in Africa and developing areas of the globe [3]. A goal to eliminate viral hepatitis was adopted by World Health Assembly for a 90% and 65% reduction in new infections and mortality respectively [4].

Sub-Saharan Africa is the second endemic area of HBV is highly endemic [5] and prevalence that is 10 times more than the Western world associated with 4% and 5.8% of deaths in South Africa and Nigeria, with the prevalence of Nigeria 12.2% among the general population [6]. North African countries including Egypt, Libya, Tunisia, Algeria, and Morocco have an intermediate rate of HBV infection ranging from 2% to 8% [7], whereas the current rate of the infection within the whole Sub-Saharan region ranges between 8 to 20% with the main transmission route is like that of Asia, that is early childhood transmission and contaminated medical equipment. 6.1% of the adult in African population is infected with HBV [8].

In Uganda, HBV infection is highly endemic, with a national prevalence of about 4.3% [9], although the distribution of the virus varies from region to region with the northern and the North-Western part of Uganda has the highest prevalence of HBV [10, 11], with high prevalence among the fishing communities of Lake Victoria [12]. All adults at risk of acquiring the virus are supposed to be screened and vaccinated against HBV as it is about 95% effective due to the development of herd immunity, hence preventing the infection and its chronic consequences from occurring [13]. In this study, we sought to establish factors influencing the prevalence of Hepatitis B viral infection among adults in Moyo district, North-Western Uganda.

## Methods

### Study Design

We used a descriptive cross-sectional study where both quantitative data collection methods and analysis were employed. Self-reported diagnosis of adults who had ever tested for HBV in the last one year, with test result forms reviewed by the researchers to confirm whether they were negative or positive with HBV. 384 samples were determined using Cochran (1963:75) and a multi-stage sampling technique was used. Bivariate and multivariate analyses were done using SPSS (20.0).

### Participant selection and inclusion criteria

Our study included all adults aged 18 -45 years who had been previously tested for HBV, having test results and were able to avail their medical record forms to the researchers to confirm whether they tested positive or negative with Hepatitis B viral infection. Respondent should have lived in the district for at least six months and consented to participate in the study.

### Data Analysis

Our data were analyzed using SPSS version 20. Chi-square statistics were computed to check for statistically significant differences in the parameters between the dependent and independent variables. Logistic regression was done to obtain strength of association between categorical dependent and independent variables and statistically significant items at this level were then analysed using multivariate logistic regression to obtain adjusted conclusions for the study.

## Results

### Demographic Characteristics of respondents

In this study 384 respondents aged 18-45 years participated, with almost an equal number of participants in all the age groups. Respondents between18-23 years were the highest in number 97(25.3%) followed by the age group 24-35 with 93(24.2%), 30-35(22.4%) and 40-45(20.1%), whereas age group 36-39 had the least number of respondents 31(8.1%). Female respondents 200(52.1%) were slightly higher than male respondents 184(47.9%). Married people had the highest number of respondents 179(46.6%) followed by those who are single 140(36.5%) with the least being Widowers 2(0.9%). In regards to education level, the majority of respondents had reached at least secondary level, 140(36.5%) secondary, tertiary 112(29.2%), the least being 27 (7.0%) were those who had never gone to school. Regarding the employment status of the respondents, the majority of those who participated in this study were adults who were engaged in subsistence farming, business and construction work 276 (71.9%) followed by health care workers 58(15.1%) (See Table 1).

**Table 1:** Demographic Characteristics of respondents.

| Variable | No. (384) | Percentage (%=100) |
| --- | --- | --- |
| Age group |  |  |
| 18-23 | 97 | 25.3 |
| 24-29 | 93 | 24.2 |
| 30-35 | 86 | 22.4 |
| 36-39 | 31 | 8.1 |
| 40-45 | 77 | 20.1 |
| Total | 384 | 100 |
| Gender |  |  |
| Male | 184 | 47.9 |
| Female | 200 | 52.1 |
| Total | 384 | 100 |
| Marital Status |  |  |
| Married | 179 | 46.6 |
| Cohabiting | 26 | 7.0 |
| Single | 140 | 36.5 |
| Divorced | 25 | 6.5 |
| Widow | 11 | 2.9 |
| Widower | 2 | 0.9 |
| Total | 384 | 100 |
| Level of Education |  |  |
| None | 27 | 7.0 |
| Primary | 105 | 27.3 |
| Secondary | 140 | 36.5 |
| Tertiary | 112 | 29.2 |
| Total | 384 | 100 |
| Employment Status |  |  |
| Health care work | 58 | 15.1 |
| Teaching/ Instructing | 27 | 7.1 |
| Barber Solon | 23 | 6.1 |
| Others Specify | 276 | 71.9 |
| Total | 384 | 100 |

### Prevalence of hepatitis B viral infection

In this study, out of the 384 respondents interviewed, 29 (7.6%, 95%CI: 5.1-10.7) had Hepatitis B viral infection (See Table 2).

**Table 2:** Prevalence of hepatitis B viral infection among adults in Moyo district, Northwestern Uganda.

| Variables | Frequency | Percentage (95% CI) |
| --- | --- | --- |
| Prevalence of Hepatitis B viral infection |  |  |
| Yes | 29 | 7.6 (5.1- 10.7) |
| No | 355 | 92.4 (89.3- 94.9) |
| **Total** | **384** | **100.00** |

### Factors influencing the prevalence of Hep B viral infection among adults in Moyo district, Northwestern Uganda

In our multivariate analysis, level of education (p=0.047*), ever had STIs (aOR=18.090, 95%CI=5.699-57.426, p=0.000*), Health facilities have equipment for screening HEP B viral infection ANC (aOR=10.762, 95%CI=1.316-88.027, p=0.027*) were statically significant in influencing the prevalence of Hepatitis B viral infection among adults, while the number of sexual partners ever had (p=0.984) was statistically not significant in influencing the prevalence of Hepatitis B viral infection.

Our univariate analysis showed that education level was significantly associated with the prevalence of Hepatitis B viral infection among adults, those who did not ever go to school, included respondents who ended in primary were less likely to be infected with Hepatitis B, respondents who ended in secondary were more likely to acquire Hepatitis B viral infection compared to those who studied up to tertiary institutions of learning. Respondents who had ever had any of STIs including HIV, Syphilis, gonorrhoea were more likely to acquire Hepatitis B viral infection compared to those who had never had any of the STIs. Those having less than one or two sexual partners were less likely to acquire Hepatitis B compared to respondents who had five and more sexual partners, moreover, respondents who had three of four sexual partners were more likely to acquire Hepatitis B. also, health facilities having screening equipment for Hepatitis B viral infection increases the prevalence of the disease among adults *(See Table 3)*.

**Table 3:** Prevalence of hepatitis B viral infection and associated factors among adults in Moyo district, North-western Uganda.

| Variables | Prevalence of HBV |  | Univariate Logistic Regression Analysis |  | Multivariate Logistic Regression Analysis |  |
| --- | --- | --- | --- | --- | --- | --- |
|  | Negative (%) | Positive (%) | uOR (95%CI) | P-Value | AOR (95%CI) | P-Value |
| **Level of Education** |  |  | **3** | **0.040\*** | **3** | **0.047\*** |
| None | 26(6.8) | 1(0.3) | 0.679(0.078- 5.8892) | 0.0726 | 0.275(0.028-2.732) | 0.270 |
| Primary | 101(26.3) | 4(1.0) | 0.70(0.192- 2.552) | 0.0589 | 0.361(0.028-1.586) | 0.177 |
| Secondary | 106 | 6 | 2.607(0.998- 6.807) | 0.050* | 1.653(0.531-5.145) | 0.386 |
| Tertiary Institutions | 122(31.8) | 18(4.7) | 1 |  |  |  |
| **Ever had STIs** |  |  |  |  |  |  |
| Yes | 261 | 4 | 17.354(5.885-51.177) | 0.000* | 18.090(5.699-57.426) | **0.000\*** |
| No | 94 | 25 | 1 |  |  |  |
| **Number of sexual partners ever had** |  |  | **4** | **0.040\*** | **4** | **0.984** |
| One (1) | 80 | 3 | 0.438(0.118-1.628) | 0.218 | 1.085(0.236-4.993) | 0.917 |
| Two (2) | 137 | 6 | 0.375(0.080-1.757) | 0.213 | 1.043(0.185-5.884) | 0.962 |
| Three (3) | 40 | 4 | 1.538(0.432-5.473) | 0.506 | 1.390(0.336-5.766) | 0.650 |
| Four (4) | 52 | 8 | 1.739(0.487-6.210) | 0.394 | 1.373(0.328-5.748) | 0.664 |
| Five (5) and above | 46 | 8 | 1 |  |  |  |
| **Health facilities have equipment for screening HEP B.** |  |  |  |  |  |  |
| Yes | 101 | 1 | 11.134(1.495-82.924) | **0.019\*** | 10.762(1.316-88.027) | **0.027\*** |
| No | 254 | 28 | 1 |  | 1 |  |

## Discussion

Our study found that the prevalence of HBV among adults in Moyo district in North-Western Uganda was 7.6%, this prevalence similar to previous studies conducted in the region [10, 11], among fishermen along with shares of lake Victoria [12] and higher than national prevalence [9]. Factors influencing this prevalence included education level which was significantly associated with the prevalence of Hepatitis B viral infection among adults, those who did not ever go to school, included respondents who ended in primary were less likely to be infected with Hepatitis B, respondents who ended in secondary were more likely to acquire Hepatitis B viral infection compared to those who studied up to tertiary institutions of learning. Similarly, study Peng *et al* showed that HBV infection was associated with low education level [14]. Also, a study conducted in Northern Ethiopia showed an association between education level and HBV infection [15]. Contrary to our study, Mustapha et al noted those adults who had reached tertiary education were less likely to test positive with HBV [6].

Our study found that respondents who had other STIs like HIV, syphilis, and gonorrhoea were more likely to acquire Hepatitis B viral infection, this means that there is a need for improved screening service among PLWHA, including vaccination [16]. Similarly, a study conducted in Nigeria noted that sexually active adults with STIs are at high risk of acquiring Hepatitis B viral infections [17]. Agrawal *et al* and Govindan et al found a lower prevalence of HBV among patients infected with syphilis [18, 19], other studies have noted a high prevalence of syphilis, HIV, HCV and Hepatitis B co-infection [20, 21, 22, 23, 24, 25, 26].

Our study found that respondents who had three or more sexual partners were more likely to acquire HBV infection. One of the major routes of HBV transmission is through sexual transmission [27]. Asaye *et al* noted that HBV infection is linked history of having multiple sexual partners [28, 29]. Contrary to the study, Adekanle *et al* noted a higher prevalence of HBV infections among respondents who had no sexual partner [30].

Health facilities having screening equipment for Hepatitis B viral infection increases the prevalence of the disease among adults. Screening and testing for HBV should be implemented in high endemic areas as it helps ascertain the burden of the disease [31, 32], HBV screening equipment helpful large sample screening [33].

### Study strength and limitations

Our study was the first study on HBV conducted in the study area, reviewed test result forms of respondents and had a large sample size. As a limitation, we were unable to test respondents to confirm the self-reporting, although HBV diagnosis was confirmed through the test forms available with some respondents, this might have reduced or even overrated HBV prevalence in the study area.

## Conclusion

We found a high (7.6%) prevalence of HBV among adults in Moyo district in North-Western Uganda compared to the national prevalence, similar to previous studies conducted in the region. The key influence of HBV prevalence included education level, ever being infected with any other STIs (HIV, Syphilis, gonorrhoea), multiple sexual partners and presence of HBV screening equipment at the health facilities. Need for more emphasis on HBV childhood immunization, screening, vaccination of adults, other preventive measures and treatment of those already infected.

## Data Availability

All relevant data are within the manuscript and its supporting information files.

## Ethical Approval

This study was approved by the International Health Sciences University Research Ethics committee (Currently Clarke International University) and administration clearance was done by Moyo District Health Office.

## Abbreviations

HBV: Hepatitis B viral Infection; STIs: Sexually Transmitted Infections; HIV: human immunodeficiency virus SPSS: Statistical package for social sciences; aOR: Adjusted Odds Ratio; CI: Confidence Interval; uOR: Unadjusted Odds Ratio.

## Acknowledgement

We appreciate International Health Sciences University (Currently, Clarke International University) for awarding the primary author with a Bachelor of Science in Public Health. We appreciate all our research assistants for the support rendered during the process of data collection.

## Authors’ Contributions

PML conceptualised and designed the study, obtained the needed data, analysed the data and interpreted results, drafted made a final copy of the manuscript. JBA participated in conceptualising and designing the study, perform critical revision of the manuscript. Both PML and JBA approved the final manuscript.

## Consent for publication

No application

